# Advancing Bloodstream Infection Prediction Using Explanable Artificial Intelligence Framework

**DOI:** 10.1101/2024.04.10.24305614

**Authors:** Rajeev Bopche, Lise Tuset Gustad, Jan Egil Afset, Birgitta Ehrnström, Jan Kristian Damås, Øystein Nytrø

**Affiliations:** Department of Computer Science, Norwegian University of Science and Technology, Trondheim, Norway; Faculty of Nursing and Health Sciences, Nord University, Levanger, Norway; Department of Medical Microbiology, St. Olavs Hospital, Trondheim University Hospital, Trondheim, Norway; Department of Clinical and Molecular Medicine, Norwegian University of Science and Technology, Trondheim, Norway; Department of Computer Science, The Arctic University of Norway, Tromsø. Norway; Department of Infectious Diseases, Clinic of Medicine, St Olavs Hospital, Trondheim, Norway; Clinic of Anaesthesia and Intensive Care, St Olavs Hospital, Trondheim University Hospital, Trondheim; Department of Medicine and Rehabilitation, Levanger Hospital, Nord-Trøndelag Hospital Trust

**Keywords:** Machine Learning, eXplainable Artificial Intelligence, Bloodstream infections, Predictive analytics, Electronic Health Records

## Abstract

Bloodstream infections (BSIs) represent a critical public health concern, primarily due to their rapid progression and severe implications such as sepsis and septic shock. This study introduces an innovative Explanable Artificial Intelligence (XAI) framework, leveraging historical electronic health records (EHRs) to enhance BSI prediction. Unlike traditional models that rely heavily on real-time clinical data, our XAI-based approach utilizes a comprehensive dataset incorporating demographic data, laboratory results, and full medical histories from St. Olavs Hospital, Trondheim, Norway, covering 35,591 patients between 2015 and 2020. We developed models to differentiate between high-risk and low-risk BSI cases effectively, optimizing healthcare resource allocation and potentially reducing healthcare costs. Our results demonstrate superior predictive accuracy, particularly the tree-based models, which significantly outperformed contemporary models in both specificity and sensitivity metrics.

**Author Summary:** In this research, we have developed a new tool that uses artificial intelligence to better predict bloodstream infections, which can lead to serious conditions like sepsis if not quickly identified and treated. It is the first of its kind framework that analyzes past health records and helps identify patients who are at high risk of infection more accurately than existing tools. Unlike existing tools our framework can be implemented at any stage of the patient trajectory and is the only framework to achieve good accuracy without the use of intimate patient features such as vital signs. This ability could enable doctors to prioritize care more pre-emptively, effectively, potentially saving lives and reducing unnecessary medical tests. Our approach is designed to be easily understood and used by both medical professionals and those with little technical expertise, making it a valuable addition to hospital systems.

## 1. INTRODUCTION

Bloodstream infections (BSIs) are a significant public health challenge, often leading to severe clinical conditions such as sepsis and septic shock, particularly when unrecognized or untreated. The rapid progression of these infections, coupled with their association with high morbidity, mortality, and healthcare costs, renders BSIs a critical challenge in clinical care[1, 2]. The accuracy of available clinical decision tools for BSI and sepsis is in need of improvement. Currently, most of them are mostly based on changes in vital signs and abnormal blood test results[3, 4]. Improved prediction can lead to more efficient allocation of resources and reduced health costs. A refined initial stratification would aid in allocating resources to patients with a high risk of a BSI and reduce needless testing of patients with low risk. BC may yield not only relevant bacteria causing disease but also growth of contaminating microbes. Ambigous culture results may be difficult to interpret and lead to clinical uncertainty, often resulting in longer hospital stay and unnecessary administration of antibiotics [5, 6]. Therefore, reduced collection of BC in patients with a low risk of BSI can lead to a higher positive predictive value of blood cultures as well as reduced use of antibiotics. Prompt and early identification of high-risk versus low-risk patients is thus imperative for adequate and effective initial handling of patients with suspected BSI, optimized allocation of healthcare resources and reduction of associated costs.

The advent of Artificial Intelligence (AI) allows innovative methodologies for BSI diagnostics, showcasing the potential to enhance or even surpass human expertise in diagnostic endeavors [7]. Despite its demonstrated efficacy, the integration of AI into clinical workflows remains limited [8, 9]. Facilitating this integration may involve leveraging AI models to augment guidelines-based Clinical Decision Support Systems (CDSS) rather than striving for the development of fully autonomous AI-based CDSS [10, 11]. Furthermore, adopting a stance grounded in Predictive, Preventive, and Personalized Medicine (PPPM) principles could refine the utilization of AI, emphasizing the analysis of historical rather than real-time data variables [12, 13]. The primary aim of this study was to develop and validate an eXplainable Artificial Intelligence (XAI) framework that utilizes historical electronic health records (EHRs) to enhance the prediction of BSIs, thus facilitating early and precise intervention. By integrating a rich dataset with advanced machine learning techniques, we sought to overcome the limitations of current real-time diagnostic methods. Our findings confirm that the XAI framework not only meets but exceeds traditional diagnostic performance, demonstrating enhanced predictive accuracy and operational efficiency in BSI management. This achievement marks a significant step forward in the application of AI in medical diagnostics, potentially transforming patient outcomes in healthcare settings.

## 2. LITERATURE REVIEW

BSI are a critical precursor to sepsis, a severe and potentially life-threatening condition. Early detection and management of BSI can significantly mitigate the risk of progressing to sepsis. In this context, leveraging AI models to predict and manage BSI presents a promising avenue. Our literature review sought to capture the breadth and heterogeneity of recent advances in machine learning ML-based models for BSI prediction. We compiled and analyzed thirty studies published in the last five years, focusing on various healthcare settings and patient demographics [14–44]. These studies predominantly focused on inpatient settings, constituting 56% (n=17) of the research, followed by emergency department (ED) settings at 23% (n=7), and ICU settings at 20% (n=6). Within the inpatient group, the studies varied, with nine examining general populations [14–22], two targeting inpatients with central venous catheters (CVC) [23, 24], and others focusing on specific patient categories such as, hemodialysis (HD) patients [26], cancer patients [27], maternity patients [28], patients with low procalcitonin levels (PCT ≤2.0 ng/ml) [29], and HIV patients [30]. Bacteremia was the primary condition under study in 24 articles, including varied focuses such as fungemia [14] and Candidemia [27, 43], while three studies aimed at predicting central line-associated bloodstream infections (CLABSIs) [33, 34, 41] and one on hospital-acquired BSI (HA-BSI) [22]. All articles reported high prediction performance (AUROC > 0.7) except for one article [20] which reported poor performance metrics. Most studies were single centered, three articles sourced their data from two hospitals [14, 16, 38], and two articles used data from multiple centres [35, 43]. The key characteristics such as, target condition, number of patients or samples, data source, prevalence, ML models, and top predictors for each study grouped by settings are shown in Table 1 on Page No. 28.

**Table 1.** Comparison of the predictors (Top 25 most influential) between the two classes.

| Feature names | Class 1 (mean)<br>(n=5288) | Class 0 (mean)<br>(n=60687) | T-Statistics | P-Value |
| --- | --- | --- | --- | --- |
| ICD_A (Infectious diseases) | 0.344175492 | 0.127045331 | -41.35067292 | 0 |
| ICD_B (Infectious diseases) | 0.270990923 | 0.098044062 | -34.86262822 | 6.9733E-264 |
| ICD_N (Kidney disease) | 0.498108926 | 0.255112298 | -31.06397543 | 2.4511E-210 |
| BILIRUBIN TOTAL (Total bilirubin) | 14.64849131 | 7.316773443 | -23.83455936 | 6.0583E-125 |
| prior_positive_blood_culture_test | 0.370083207 | 0.178852143 | -20.33423563 | 1.22442E-91 |
| ICD_J (Influenza, pneumonia etc) | 0.307677761 | 0.488885593 | 16.95136079 | 2.5746E-64 |
| ICD_R (abnormal symptoms and findings) | 0.298411498 | 0.188771895 | -16.06835215 | 5.48158E-58 |
| Age (at the time of BC) | 68.87821483 | 65.07332707 | -14.51708417 | 1.11892E-47 |
| KREATININE (Creatinine) | 99.42019885 | 82.73778714 | -13.30695014 | 2.42534E-40 |
| CRP | 110.8463237 | 94.62243471 | -12.20533489 | 3.21434E-34 |
| ICD_A_aggregate | 0.797655068 | 0.557236311 | -11.29534135 | 1.47299E-29 |
| LEUKOCYTTER (Leukocytes) | 11.57262042 | 10.27756163 | -10.07214858 | 7.68314E-24 |
| procedure_G_aggregate | 0.996596067 | 1.48710597 | 9.453454788 | 3.3812E-21 |
| ICD_U (Unsure etiology and antimicrobial resistance) | 0.018910741 | 0.006953713 | -9.34670256 | 9.3135E-21 |
| ICD_J_aggregate | 1.876134644 | 2.99597937 | 9.271600994 | 1.88697E-20 |
| procedure_K (Procedures on the ears) | 0.073940998 | 0.038377247 | -8.468324845 | 2.5398E-17 |
| TROMBOCYTTER (Thrombocytes) | 156.5213799 | 174.202004 | 8.409629616 | 4.20742E-17 |
| ICD_K (Gastrointestinal diseases) | 0.234871407 | 0.179527741 | -8.148452108 | 3.75005E-16 |
| procedure_J (Procedures on endocrine glands) | 0.184379728 | 0.127407847 | -6.851217934 | 7.38635E-12 |
| prior_positive_edta | 0.142965204 | 0.254074184 | 6.204949673 | 5.50384E-10 |
| ICD_M (Musculoskeletal disease) | 0.067133132 | 0.092490978 | 5.438316649 | 5.39768E-08 |
| ICD_B_aggregate | 0.752458396 | 0.554468008 | -5.194454856 | 2.06E-07 |
| ICD_R_aggregate | 2.291603631 | 2.686555605 | 5.06204369 | 4.15899E-07 |
| Explicitsepsis (Sepsis episode) | 0.006807867 | 0.002751825 | -5.034421936 | 4.80552E-07 |
| LOS (Length of stay till BC tests) | 8.751426185 | 8.013890262 | -4.578791274 | 4.68523E-06 |

Among studies with study design of inpatient settings, Bhavani et al. (2020) used EHRs from two academic tertiary medical centers between 2007 and 2018. Data types included Demographic data, International Classification of Diseases billing codes, clinician orders, blood culture results, vital signs, nursing assessments, and laboratory values [14]. The LR and GBM models were developed. GBM models demonstrated superior performance in predicting bacteremia and fungemia with area under the receiver operating characteristic curve (AUC) scores of 0.78 for bacteremia and 0.88 for fungemia. Lee et al. (2019) investigated the early detection of bacteraemia using an artificial neural network (ANN) model, specifically a multi-layer perceptron (MLP). The study utilized data from 13,402 patients at Gangnam Severance Hospital in South Korea, identifying 1,260 episodes of bacteraemia from blood cultures [15]. Data included 20 clinical variables such as vital signs and various laboratory data. The study highlighted the effectiveness of MLP models, showing remarkable sensitivity in identifying bacteraemia episodes based on a well-curated set of clinical variables. Lee et al. (2022) expanded on earlier research, utilizing an extensive dataset, also from Gangnam Severance Hospital, consisting of a larger patient sample size and covering more recent patient admissions from 2013 to 2018 [16]. This study employed enhanced ANN models alongside other ML techniques like Random Forest (RF) and Support Vector Machines (SVM) to predict bacteraemia from clinical and laboratory data. The study by Mahmoud et al. (2021) developed a predictive model for bacteremia using data from 7,157 adult patients admitted to King Abdulaziz Medical City in Riyadh. This retrospective cohort study utilized EHRs from July 2017 to July 2019 to analyze 36,405 blood culture tests [20]. The dataset included demographic details, clinical variables such as vital signs (e.g., temperature, heart rate, blood pressure), and laboratory test results (e.g., white blood cell count, platelet count, creatinine level, lactic acid level, C-reactive protein (CRP), and procalcitonin levels). Several machine learning models were employed to determine the best predictor of positive blood cultures, including Neural Networks (NN), Random Forest (RF), Logistic Regression (LR), Decision Trees (DT), Naive Bayes (NB), and Support Vector Machines (SVM) with a Radial Basis Function (RBF) kernel. The study by Garnice et al. (2021) utilized a dataset from the Hospital Universitario de Fuenlabrada in Madrid, comprising 4,357 patients with 117 features per patient [21]. The features included patient demographics, medical history, clinical analysis, comorbidities, and the results of blood cultures, differentiated into cases of bacteraemia and no bacteraemia. Three supervised machine learning classifiers were implemented: Support Vector Machine (SVM), Random Forest (RF), and K-Nearest Neighbours (KNN). Each classifier was employed in two scenarios: using only the features available at the time of blood extraction, and a second scenario that included additional features revealed during the blood culture process. These studies highlight the potential of ML to enhance diagnostic accuracy and patient outcomes in detecting critical infections.

Among studies in the ED settings, Choi et al. (2022) developed ML models to predict bacteremia in emergency department (ED) settings at Seoul National University Hospital [31]. The study used data from 24,768 adult patients collected between 2016 and 2018. The models utilized demographic information, chief complaints, vital signs, and laboratory data collected during ED triage and disposition. Two primary models were developed: the Triage XGB model and the Disposition XGB model. In a subsequent study Choi et al. (2023) aimed to refine the predictive accuracy of ED triage-based bacteremia identification using an advanced ensemble of ML techniques. The study analyzed data from over 30,000 ED visits, employing a wide array of clinical inputs including detailed symptom descriptions, vital signs, and initial lab results [32]. The developed model incorporated a Gradient Boosting Machine (GBM) framework that effectively integrated the diverse dataset to predict bacteremia risk. The study by Schinkel et al. (2022) harnessed data from EHRs of 44,123 unique emergency department visits across four hospitals: Amsterdam UMC, Zaans Medical Center, and Beth Israel Deaconess Medical Center, covering the period from 2011 to 2021 [35]. The model employed was a hybrid of LR and (eXtreme Gradient Boosting) XGBoost, with the latter outperforming in predictive accuracy. The data utilized included patient demographics, vital sign measurements, and common laboratory results such as temperature, creatinine, and C-reactive protein, which were identified as the most influential predictors. This predictive model was integrated into the VUMC’s EHRs system for real-time prospective evaluation, affirming its practical utility by potentially reducing unnecessary blood culture analyses by at least 30%. The study by Boerman et al. (2022) utilized a single-center, retrospective observational design, the study encompassed data from 51,399 ED visits at the Amsterdam UMC from September 2018 to June 2020 [33]. Data included demographics, vital signs, laboratory and radiology results, and medications administered during ED visits. The study employed two predictive models: a LR model and a gradient boosted tree (GBT) model, both of which demonstrated good predictive performance with an area under the receiver operating characteristic curve (AUROC) of approximately 0.77 and 0.78 respectively. Notably, the gradient boosted tree model was optimized to predict 69% of blood cultures as negative, with a negative predictive value exceeding 94%, indicating its utility in potentially reducing unnecessary blood cultures and associated healthcare costs. The models harnessed a comprehensive array of features, including commonly available clinical data such as C-reactive protein levels and white cell counts, to predict the likelihood of bacteraemia. These studies illustrate how integrating ML models into ED workflows can improve the speed and accuracy of BSI detection, potentially reducing unnecessary interventions and optimizing resource allocation. However, all the model focused on recent data pertaining to the patients and none of the study utilized the predictors from the complete medical history of their patients, apart from demographic and information on co-morbidities. In our previous work through innovative feature engineering from historical medical records and employing an array of machine learning classifiers, we showcased the efficacy of eXtreme Gradient Boosting (XGBoost) model, in predicting 30-day mortality using EHR trajectory features [56].

## 2. MATERIALS AND METHODS

### 2.1. Source of data

EHRs provide a longitudinal perspective of patients’ interactions with hospital service. In Norway, with predominantly public specialist healthcare, patients often have long and continuous histories within one hospital’s records. This study harnessed EHRs from St. Olavs University Hospital, Trondheim, Norway, encompassing 35,591 patients with suspected bloodstream infections (BSIs) identified via physician-initiated blood cultures between 2015 and 2020. The EHRs encompassed, curated data from the inception of electronic records in 1999 until 2020, exclusively included hospital care episodes (excluding primary care and other specialist care episodes), Intensive Care Unit (ICU) admission details, microbiology test results, laboratory test results and patient demographics comprising of gender, date of birth, and date of death. Diagnoses and Procedures within these records were classified using the International Classification of Diseases, 10th Revision (ICD-10), facilitating standardized disease identification critical for the analytical models. This study adhered to the ‘transparent reporting of a multivariable prediction model for individual prognosis or diagnosis (TRIPOD) [46]. For ethical considerations the EHRs were de-identified and accessed through a private cloud computing platform.

### 2.2. XBSI Framework

The XBSI framework aggregated various data types from the raw EHRs including demographics, laboratory tests including microbiology tests, discharge summaries, and ICU admissions, as depicted in Figure 1. This dataset underwent preprocessing, event log creation for sequential ML models, and feature engineering for static ML models before being transformed and scaled to facilitate the respective model development pipelines. Following subsections details of the various steps in the XBSI framework.

**Figure 1.**
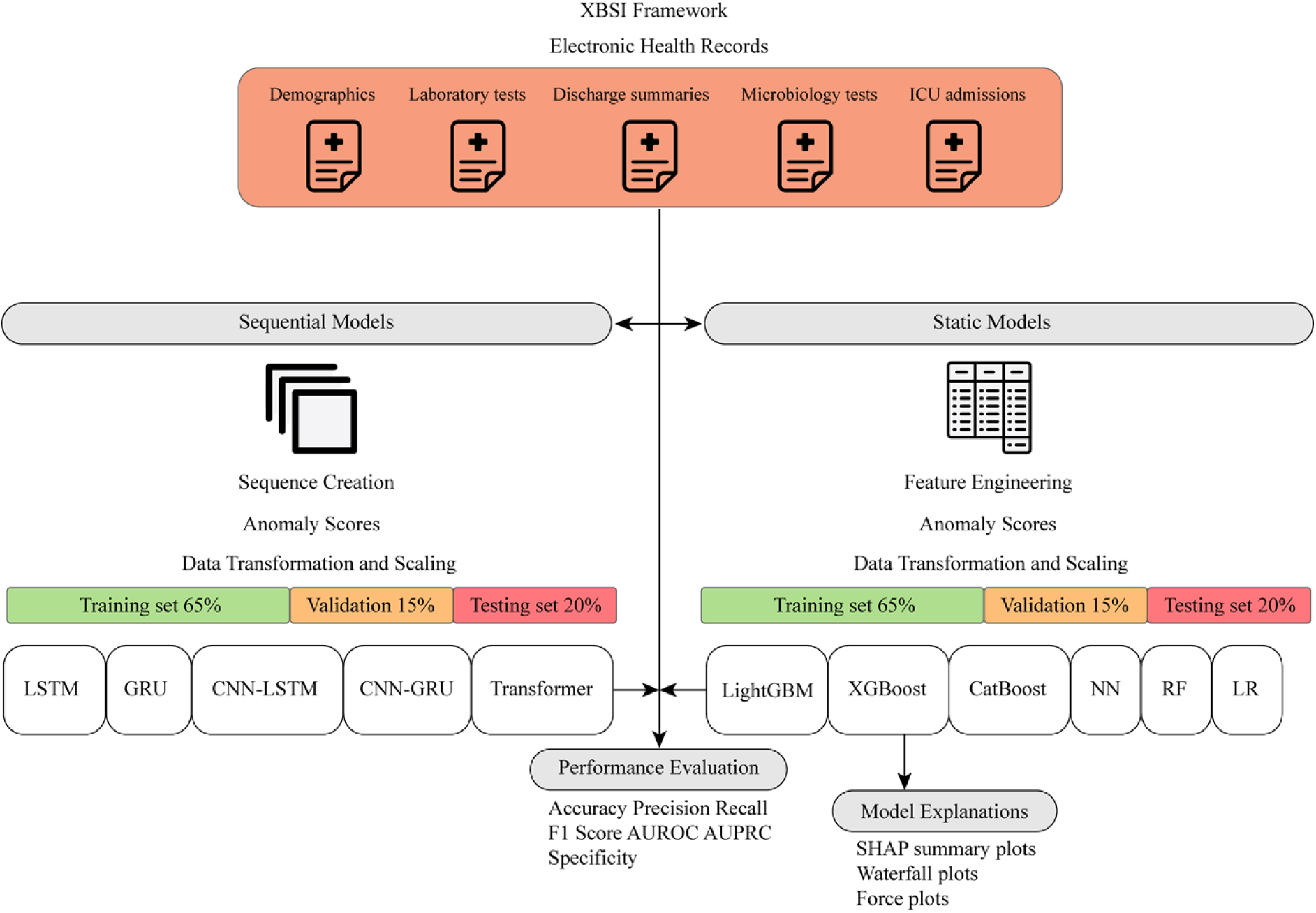
Schematic Overview of the XBSI framework. The diagram illustrates the XBSI framework applied in the study, starting with the extraction of electronic health records (EHRs), which include demographics, laboratory tests, discharge summaries, microbiology tests, and ICU stays. The workflow bifurcates into two parallel processes: Sequence Creation and Feature Engineering, both incorporating Anomaly Scores derived from the data. Subsequent steps include Data Transformation and Scaling, with the data split into training (at 65% and 80%, respectively), validation (at 15%), and testing (at 20%) subsets. The bottom layer of the workflow depicts the range of ML models, the performance metrics used to assess them, and the model explanations generated to study the feature importances.

#### 2.2.1. Data Preprocessing and Transformation

Utilizing Python libraries such as Pandas and NumPy, the CSV files comprising raw EHRs, were loaded into dataframes for further processing. The discharge summaries required several data cleaning steps to ensure quality and relevance of the data: Relevant patient information such as identifiers, admission and discharge times, and diagnostic codes were retained. Instances of missing identifiers were addressed by replacing empty strings with NaN values and subsequently removing these records. Data were organized by patient identifier and admission/discharge times to maintain coherent episode tracking. Non-standard characters within diagnostic codes, such as semicolons and commas, were standardized to spaces, and any duplicates were removed. The timestamps were converted into datetime format, facilitating the calculation of the length of stay in hours for each episode. The duration of each ICU stay was calculated in hours, along with the total count of each type of hospital admissions per patient. The request dates in both laboratory test results and microbiology test results were standardized to datetime objects and used to create eventlogs of tests per patient. Specialized functions were created to clean the laboratory an microbiology test table entries. This function performed tasks to remove any non-numeric characters, which could represent encoding errors or artifacts from data entry. It standardized decimal point characters by replacing commas with periods, which is necessary for consistent numerical representation across different regions that may use varying formats for decimal points. The microbiology test table was filtered to identify suspected BSI episodes, with an emphasis on blood culture tests. The results column was processed to standardize and clean the values, categorizing them as ‘positive’, ‘negative’, or ‘contaminant’ based on the results column. For each patient ID, four event logs were created, from discharge summaries, ICU admissions, laboratory tests and microbiology tests, following subsection describes the event logs.

#### 2.2.2. Event log description

*Discharge Summaries Event Log:* Captures patient discharge information, including admission and discharge times, diagnostic and procedural codes, urgency and care level code.

*ICU Admissions Event Log:* Records details of each ICU stay, including the duration in hours and the total count of ICU admissions per patient and total length of ICU stays per patient.

*Laboratory Tests Event Log:* Includes results of various laboratory tests standardized and organized chronologically for each patient.

*Microbiology Tests Event Log:* Consists of microbiology test results, grouped by collection sample type, and categorizes them based on outcomes such as ‘positive’, ‘negative’, or ‘contaminant’. Groups of microbiology tests categorized by collection sample type are given in Supplementary List 1.

#### 2.2.3. Sequence creation

The sequence creation process was implemented using the create_sequences function, designed to compile a comprehensive view of a patient’s medical history over their entire recorded history. The create_sequences function systematically constructs a timeline of medical events for each patient, for each BC test. The medical events were merged from the four event logs. The filtered event logs are merged into a single dataframe, ensuring no information is lost. This step involves an outer join on patient ID and date, maintaining all records from each event log.

#### 2.2.4. Feature engineering

This approach involved creating a dataset with attributes derived from ICD codes, procedure codes, laboratory, and microbiology test results. Laboratory test results were organized using pivot tables, ensuring a structured format for analysis. Tests such as ‘bilirubin’ (total, conjugated, and unconjugated), ‘C-Reactive Protein’ (CRP), and ‘lactate’ (various measurements) were included, alongside white blood cell count (leukocytes), platelet count (thrombocytes), and blood gas measurements (pH, PO2). Similarly, microbiology test results were consolidated to reflect various sample types such as blood, urine, and other fluids, employing a dictionary mapping to streamline similar types. The resulting pivot table included columns for diverse samples, ranging from ‘blood culture test’ to ‘urine’, ‘feces’, and ‘nasal swabs’. A function was created to determine the aggregated result of blood culture tests for each patient group, considering the possibility of concurrent positive and contaminant results. This aggregation provided a comprehensive view of the infection status per patient. Moreover, the history of prior positive, negative, and contaminant results was calculated and added to the dataset, offering a valuable perspective on the patient‘s previous encounters with BSIs. Comorbidities were extracted and processed to identify unique diseases from patient records. New columns were created for each disease, and the counts were updated based on patient history. Finally, aggregate columns were added for both ICD and procedure codes to calculate cumulative sums. This method enabled the capturing of the cumulative history of medical conditions and procedures for each patient. All the NaN values representing absence of a condition or measurement were filled with zero and “0” is not interpreted as a value.

#### 2.2.5 Model Development

This section outlines the development and evaluation of ML models. The process bifurcates into the sequence creation and feature engineering pipelines for the temporal and static ML pipelines respectively. An Isolation Forest model was implemented to detect anomalies within the data. This model was trained exclusively on normal data (negative BC episodes) and then used to compute anomaly scores for the training, validation, and test sets. These scores were normalized and appended to the original dataset to serve as additional features, enhancing the model’s ability to distinguish between normal and anomalous patterns. Given the sequential and tabular nature of the input data for the respective ML pipelines, the necessary transformation steps were employed to prepare it for the corresponding learning algorithms. The data was reshaped and scaled using the StandardScaler from sklearn.preprocessing. This normalization step is crucial for models that are sensitive to the scale of input features. After scaling, the data was reshaped back to its original form, ensuring compatibility with the machine learning models used. Temporal ML models included Long Short-Term Memory (LSTM), Gated Recurrent Unit (GRU), Convolutional Neural Network-Long Short-Term Memory (CNN-LSTM), Convolutional Neural Network-Gated Recurrent Unit (CNN-GRU), and Transformer models. These models were trained on sequences of medical events to capture temporal dependencies within the data. On the other hand, the static ML models, which treat data points as independent and identically distributed, comprised of Light Gradient Boosting Machine (LightGBM), eXtreme Gradient Boosting (XGBoost), Categorical Boosting (CatBoost), Artificial Neural Network (ANN), Random Forest (RF), and Logistic Regression (LR). Detailed information of each ML model is given in Supplementary Methods, Section 1.1., 1.2.

#### 2.4.6 Model Validation

For cross validation, we chose a 5-fold cross-validation scheme. This technique involves dividing the entire dataset into ‘k’ equally sized subsets or folds. The model is then trained on ‘k-1’ folds and tested on the remaining fold. This process is repeated ‘k’ times, each time with a different fold as the validation set. In addition to cross-validation, we also validated our model on an independent test set, which was separated from the dataset at the outset and not used during the training phase. The test set comprised of the most recent 20% of the BC episodes while the earliest 80% of BC episodes were assigned as training cohort. Further, the training data was divided into a smaller training set and a validation set, with 15% of the training data allocated for validation. This additional split allowed for the tuning of hyperparameters and the assessment of the model’s performance during training. Each model was evaluated using a variety of metrics to evaluate model performance, including accuracy, precision, recall, F1-score, specificity, the Area Under Precision-Recall (PR) Curve (AUPRC) and the Area Under the Receiver Operating Characteristic (AUROC). Detailed information on each performance metric is given in Supplementary Methods, Section 1.3.

#### 2.4.7 Model Explanations

For interpreting the outputs of our ML models, we employ both global and local explanation methods to provide comprehensive insights into feature importances which guide clinical decision-making. To evaluate the overall impact of features across all predictions, we generate SHAP (SHapley Additive exPlanations) summary plots [55]. These plots aggregate SHAP values to illustrate the average influence of each feature on the model output, ranked by significance. This method, rooted in game theory, decomposes a prediction into the contribution of each feature, providing a transparent view into the predictive process. SHAP’s suitability for tree-based models was enhanced by its efficient computation of exact SHAP values using the TreeExplainer algorithm [57], which significantly reduced computational complexity by exploiting the structural properties of decision trees [58]. By integrating SHAP summary plots, we can discern which features are most instrumental globally in predicting bloodstream infections, facilitating an understanding of model behavior over the entire dataset On an individual prediction level, we utilize waterfall plots and force plots to detail the contribution of each feature to specific predictive outcomes. Waterfall plots provide a step-by-step breakdown of how each feature’s value contributes to the final prediction, starting from the base value (the average model output across all data points) and adding the effect of each feature sequentially. This visualization helps in understanding the decision-making process for individual predictions, which is crucial for clinical validation and personalized patient insights. Force plots, another type of local interpretability tool, display how each feature’s value pushes the model’s prediction higher or lower, which is particularly useful for individual patient assessments. These plots highlight the positive or negative contribution of each feature towards the final prediction, allowing healthcare providers to grasp the underlying reasons for a model’s decision on a case-by-case basis.

### 2.2. Participants and Outcome

All adults (aged ≥ 18 years) who had at least one BC episode during their hospital stay or visit, which was identified and ordered by a physician on the grounds of suspicion of a BSI. The primary outcome was whether a blood culture episode was positive or negative for bacteremia. A BC episode was defined as a distinct nonoverlapping 24-hour period in which one or more BC tests were ordered [44]. If one or more results within a BC episode were positive, then the BC episode was considered positive. BC results with contaminants were considered negative results [45]. The list of microbes considered as contaminants is given in Supplementary Table 6.

### 2.3. Predictors

We used data available till the date of a BC episode for training the ML prediction models. The predictors included age, sex, results of the recent laboratory test values, previous positive microbiology tests, count of co-morbidities, ICD and procedural codes, and total ICU stays. The most common laboratory tests were Bilirubin, C-Reactive Protein (CRP), Creatinine, Leukocytes, and Thrombocytes. The counts of prior positive results of microbiology tests grouped by their collected sample type were calculated and used as predictors of previous history of infections (Supplementary List 1). From the medical history the predictors included, counts of the occurrences of different ICD-10 diagnostic and the Nordic Medico-Statistical Committee (NOMESCO) Classification of Surgical Procedures (NCSP), codes and Classification of Medical Procedures (NCMP) codes, were classified according to the initial character (alphabetic), corresponding to the various chapters in of the ICD-10. Each patient record was expanded with new columns for the counts in the recent and complete history of the corresponding to each character, incrementing the count for each instance where a character led the code. For calculation of diagnostic and procedural codes counts in the recent episode, the current episode and any admissions or visits within one month of the BC test date were merged into the current medical episode. The LOS feature stored the total length of stay in the hospital for the current episode till the time of BC. The total LOS feature stored the value of the cumulative hospital length of stay (LOS) per patients. The description of each predictor and its mean across the dataset is given in the Supplementary Table 4. Further stratification was conducted by categorizing ICD codes into disease groups pertinent to clinical significance, such as ‘explicit sepsis’, ‘infection’, and ‘organ dysfunction’, among others. The table depicting the ICD codes selected for different disease groups is given in Supplementary Table 5.

### 3.4. **Prediction task modeling**

We undertook prediction modeling using two distinct datasets: a static model dataset, *X*, and a sequential model dataset, *Y*. The static dataset, *X*, encompasses labelled aggregated, patient-specific information available till the day of BC episode and labels for. Using *X*, we trained static models *M*_sta_ to predict the likelihood of a positive BC episode. the sequential dataset, *Y*, contains labelled sequences of medical events per patient compiled from the eventlogs for each BC test. The sequential models *M*_seq_ is developed using *Y* to predict the likelihood of positive BC episode.

### 2.5. Statistical Analysis

The mean values for the significant features across the two classes were computed, providing an understanding of how each feature varies with the BC episode results. Statistical analysis was performed using independent t-tests to compare the means of each feature between two independent groups labeled by blood culture test results [47]. A Pearson correlation matrix was constructed for the significant features to examine the strength and directionality of the relationships between them [48]. The resulting coefficients was visualized using a heatmap.

## 3. RESULTS

### 3.1. Patient Characteristics

There were a total of 72,495 BC episodes in the dataset. Following the exclusion of pediatrics and outpatient BC episodes 65,975 adult inpatient BC episodes were included in the analysis. Of the BC episodes, 5,288 (8%) were classified as positives. Please see the flow chart provided in Figure 2. The differences in the mean values for all feature across the two classes along with the T-statistic and p-value is given in Table (Supplementary Results, Supplementary Table 2). The top 25 most significant features and the comparison between their mean values across the two classes, along with the T-statistic and p-value are given as Table 1. There were significant differences between the positive and negative blood culture groups regarding the occurrence of ICD codes starting with A, B, and N in the medical records. The ICD Chapter I (A00-B99) describes certain infectious diseases and parasitic diseases and ICD Chapter XIV (N00-N99) concerns kidney and urinary disorders. From laboratory markers, bilirubin, creatinine, and CRP showed the most significant differences between the groups. The positive BC group also had higher mean age, higher total length of stay (LOS) till the time of BC, shorter time to the last, most recent episode, and a higher number of previous positive BC test results. The correlation matrix heatmap depicts a visual representation of the strength and directionality of correlations between various clinical features within the EPRs, highlighting the complex interdependencies relevant to BSI prediction, and is presented in Supplementary Figure 5. The top three most correlated coefficients among significant features is given in Supplementary Table 3. The list of all the features, their description, and the mean value across the dataset is given in Supplementary Table 4.

**Figure 2.**
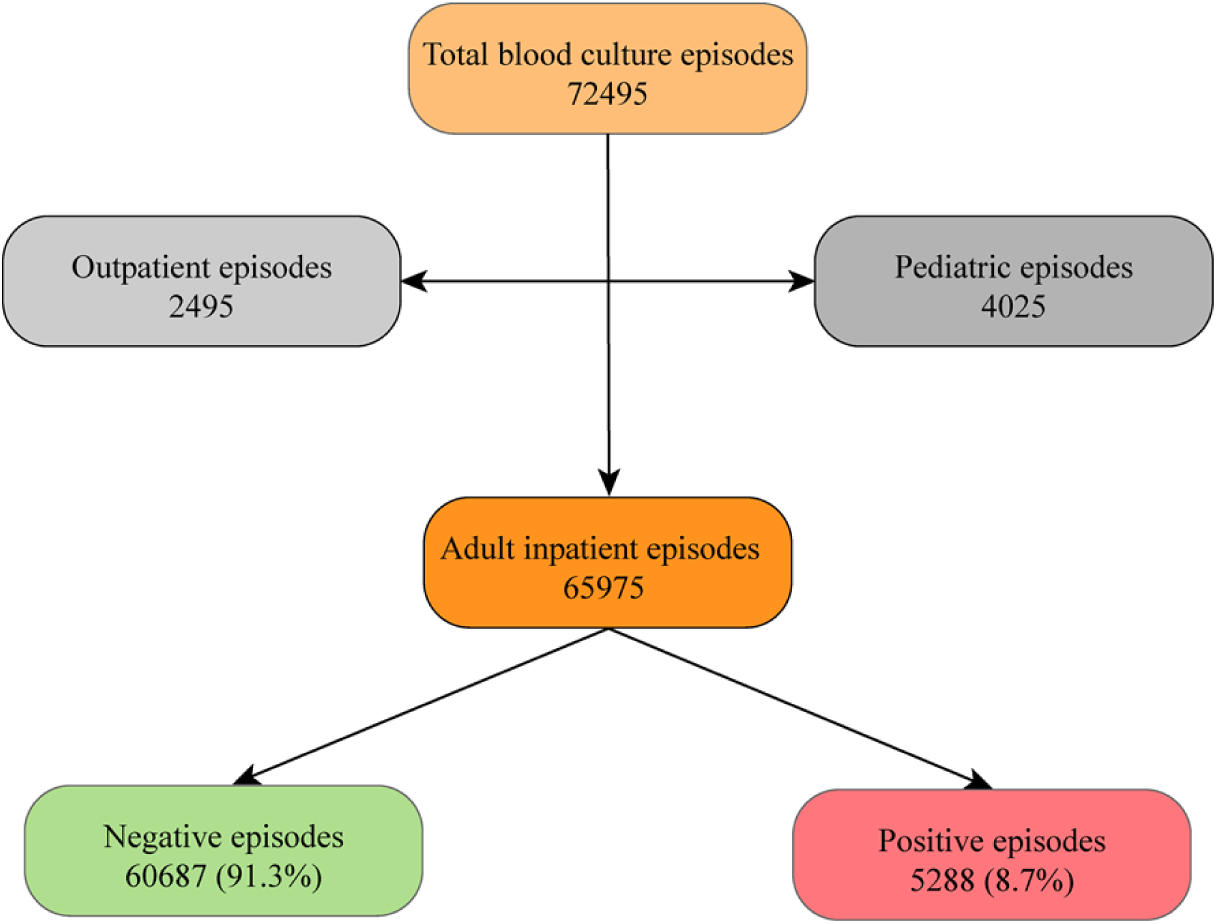
Flow chart. The flowchart depicts the categorization of the BC episodes.

### 3.2. Model Performance

Comparative performance metrics of the ML Models is given as Table 2. The Prediction performance for the ML models was evaluated using the testing set. For the sequential models, the CNN-LSTM model achieved the highest AUC of 0.7182 (95% CI 0.7064-0.7300) and AUPRC of 0.2752. The LSTM model had the highest Accuracy of 0.8955 and Precision of 0.3084. The Transformer model demonstrated the highest Recall/sensitivity of 0.4688. The GRU model gave the highest F1-score of 0.3349. In contrast, the static models outperformed the sequential models, with the CatBoost model exhibiting the highest AUC of 0.8167 (95% CI 0.8049-0.8285) and F1-score of 0.4132. The RF model showcased the highest AUC of 0.8410 (95% CI 0.8292-0.8528). However, it is notable that both the RF and NN models, with their high accuracy and specificity scored least in terms of recall. On the other hand, the LR model achieved the highest recall, in contrast to the lower performance across other metrics. The balanced performance of both XGBoost and LightGBM models across all metrics underscores the robustness of tree-based models compared to their sequential counterparts.

**Table 2.** Comparative Performance Metrics of the ML Models.

| Model | AUC (95% CI) | Accuracy | Precision | Recall | F1<br>Score | AUPRC | Specificity |
| --- | --- | --- | --- | --- | --- | --- | --- |
| *Sequential Models* |  |  |  |  |  |  |  |
| LSTM | **0.6961 (0.6843-0.7079)** | 0.8955 | 0.3084 | 0.2767 | 0.2913 | 0.2503 | 0.9422 |
| GRU | **0.7155 (0.7037-0.7273)** | 0.8652 | 0.2862 | 0.4063 | 0.3349 | 0.2599 | 0.8985 |
| CNN-LSTM | **0.7182 (0.7064-0.7300)** | 0.8826 | 0.3048 | 0.3529 | 0.3265 | 0.2752 | 0.9224 |
| CNN-GRU | **0.7020 (0.6902-0.7138)** | 0.8697 | 0.2735 | 0.3475 | 0.3054 | 0.2546 | 0.9093 |
| Transformer | **0.7015 (0.6897-0.7133)** | 0.8119 | 0.2829 | 0.4688 | 0.3047 | 0.2307 | 0.8351 |
| *Static Models* |  |  |  |  |  |  |  |
| XGBoost | **0.7951 (0.7833-0.8069)** | 0.8591 | 0.3533 | 0.5680 | 0.4025 | 0.3942 | 0.8876 |
| LightGBM | **0.8105 (0.7987-0.8223)** | 0.8103 | 0.3441 | 0.6628 | 0.3737 | 0.3987 | 0.8259 |
| CatBoost | **0.8167 (0.8049-0.8285)** | 0.8529 | 0.3523 | 0.6274 | 0.4132 | 0.4203 | 0.8750 |
| NN | **0.7693 (0.7575-0.7811)** | 0.9261 | 0.5126 | 0.2886 | 0.3634 | 0.3618 | 0.9775 |
| LR | **0.7751 (0.7633-0.7869)** | 0.7534 | 0.2462 | 0.6812 | 0.3219 | 0.2861 | 0.7627 |
| RF | **0.8410 (0.8292-0.8528)** | 0.9218 | 0.8510 | 0.2201 | 0.1990 | 0.4545 | 0.9882 |

### 3.3. Global Feature Importances

The SHAP summary plots in Figure 3. reveal the feature importances derived for the XGBoost model. The bar plot indicates that the count of ICD codes starting with letters A, B, and N, bilirubin (BILIRUBIN TOTAL), creatinine (KREATININ), leukocytes (LEUKOCYTTER), thrombocytes (TROMBOCYTTER), age, number of previous positive blood culture results, and the LOS of current or recent hospital episode were the top ten most important features for predicting positive blood culture. The beeswarm plot further elucidates the directional impact of each feature’s value on the model’s predictions. Higher values of bilirubin, creatinine, and leukocytes (as indicated by red dots on the right side of the x-axis) lead to a greater increase in the model’s prediction values, potentially signifying a higher risk whereas a lower value for thrombocytes signifies a higher risk of bloodstream infection. Other significant factors towards a positive prediction were higher cumulative hospital length of stay (total_los), lower time to the recent episodes, and higher CRP.

**Figure 3.**
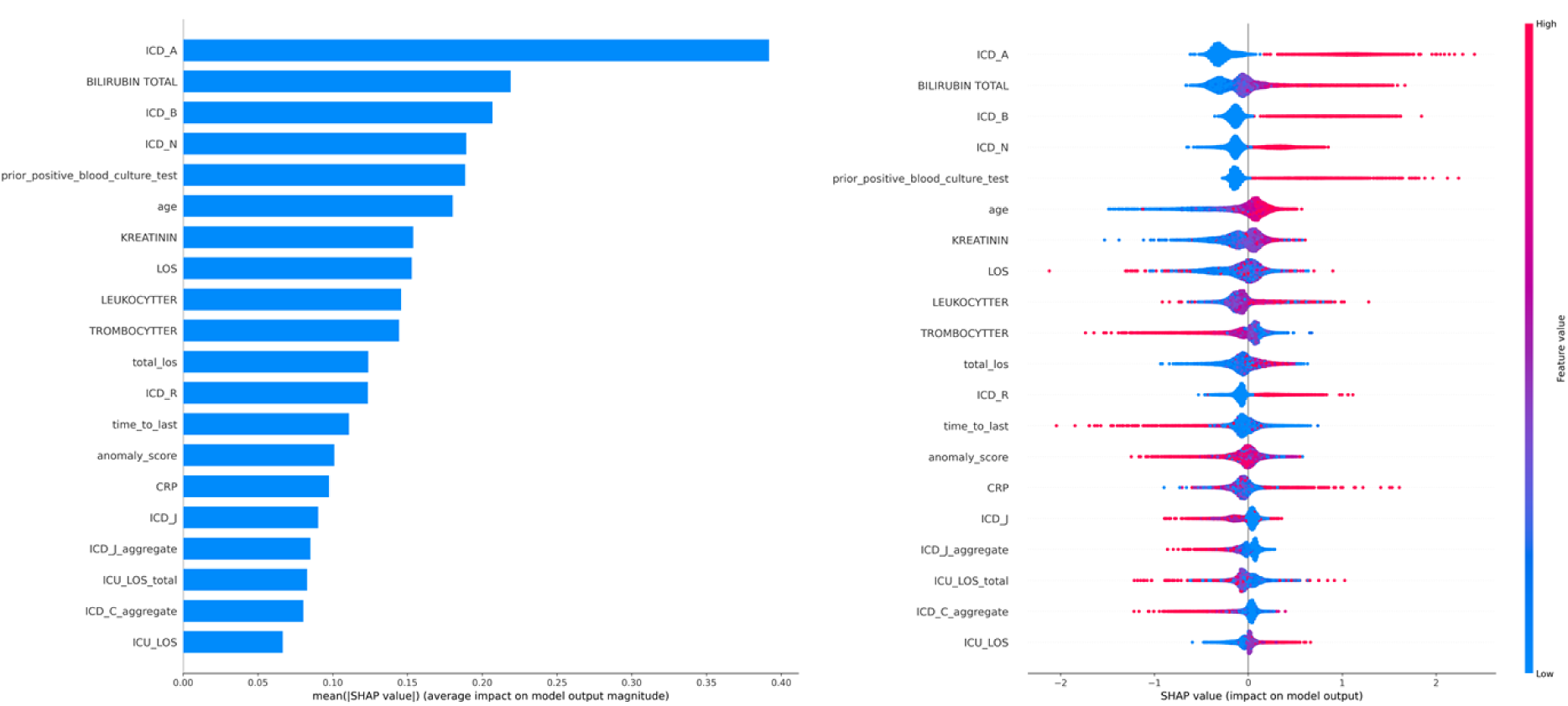
SHAP Summary Plots for XGBoost Model Feature Importance. Bar plot on the left illustrating the global feature importance ranked by the sum of SHAP values across all samples. On the right is the Beeswarm plot detailing the individual SHAP values for each feature and their impact on the model’s output.

### 3.4. Local Feature Importances

The figure 4. on Page No. 32. Details the waterfall and force plots for the first three prediction tasks. In the first prediction task, the feature ICD_A (representing infectious and parasitic diseases) has a significantly high positive SHAP value, indicating that an increase in counts of ICD-10 Chapter I codes strongly sways the model towards predicting a bloodstream infection. The ‘anomaly_score’ feature contributes negatively, suggesting that anomalies detected by the model are not predictive of the outcome. Similarly, creatinine level and ICD_N (genitourinary diseases) show negative SHAP values, implying that higher levels of creatinine and the presence of genitourinary diseases are linked to a lower probability of bloodstream infection in this instance. For the second prediction, counts of prior positive BC results has the most substantial positive impact, which aligns with clinical reasoning that past positive tests could indicate a higher risk for current infection. Additionally, ICD_B (infectious and parasitic diseases) and ICD_R has a positive effect, and total bilirubin has a positive contribution, suggesting high risk of BSI. In the third prediction, similar to the second prediction, counts of prior positive BC results again shows a positive influence. Additionally, ‘ICD_C_aggregate’ (aggregate count of cancer-related codes) exerts a negative influence on the prediction outcome, whereas ‘procedure_W_aggregate’ (count of procedures on female reproductive organs in the history) is associated with an increase in the risk of BSI.

**Figure 4.**
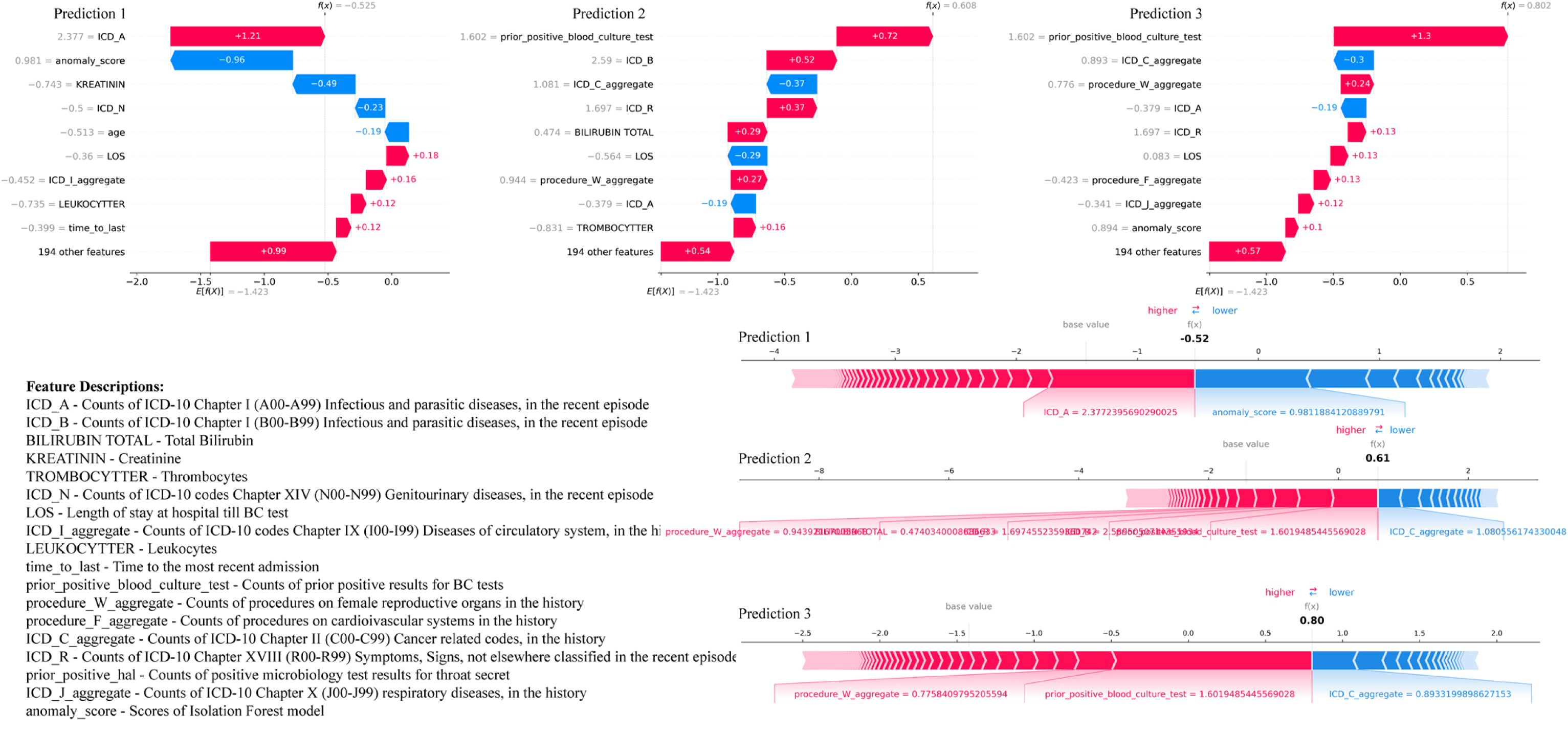
Waterfall and Force plots for the first three predictions from the test set.

## 4. DISCUSSIONS

In this study we presented the XBSI framework for early prediction of BSI in the hospitals. For the sequential models, the AUC ranged from 0.6961 to 0.7182, indicating a modest discriminatory capacity, with CNN-LSTM marginally leading in performance. Notably, the Transformer model demonstrated the highest sensitivity 0.4688. In contrast, static models depicted better AUC values ranging from 0.7751 to 0.8410. Our findings underscore the superior performance of static ML models, particularly those employing tree-based algorithms such as CatBoost, which achieved a notable AUC of 0.8167 [52]. The statistical analysis and global and local SHAP value interpretations reveal that the count of diagnostic and procedural ICD codes and routine laboratory significantly contribute to the model’s predictions. These findings align with existing literature, underscoring the clinical relevance of laboratory tests and medical history for early BSI prediction [14, 15, 16, 20, 21, 51]. The demonstrated efficacy of our XBSI framework highlights the predictive capability of combining medical history with routine laboratory tests for early BSI detection. The modular design of our framework reinforces robustness to all types of medical data and emphasize the utility of integrating diagnostic and procedural ICD codes into predictive models. This approach allows for a nuanced understanding of patient profiles, which is pivotal for implementing predictive healthcare solutions effectively.

Comparing the recent studies on predicting BSI in hospital settings, Bhavani et al. (2020) utilized LR and GBM models to predict bacteremia and fungemia from EHRs, achieving AUCs of 0.73 and 0.88 respectively. The key predictors identified in the study were time from admission to BC, temperature, age, heart rate, prior bacteremia/fungemia, white blood cells (WBC), blood urea nitrogen (BUN), glucose, diastolic blood pressure (DBP), and systolic blood pressure (SBP) [14]. Lee et al. (2019) compared multiple ML algorithms, including MLP, SVM, and RF, achieving their best AUC of 0.732 with the RF model [15]. Our study surpasses this benchmark, suggesting that the sequential analysis of patient data may capture the temporal dynamics of BSI development more effectively than models relying solely on static point-in-time data. The key predictors reported by the study included, alkaline phosphate (ALP), platelet, maximum body temperature, SBP, WBC, CRP, ICU stay, hospital day to blood culture, age, heart rate, prothrombin time, and albumin. In a subsequent study, Lee et al. (2022) further explored MLP, RF, and XGBoost among others, focusing on their application over a long-term dataset. Their models achieved AUCs of 0.762 for MLP and 0.758 for RF in the 12-hour data group, which are slightly lower than our CatBoost model’s performance. the key predictors in this study were monocyte, platelet, hospital stay, neutrophil, total bilirubin, BUN, albumin, ALP, WBC, CRP, creatinine, pulse rate, and chloride [16]. The study by Mahmoud et al. used data from a tertiary care center comprising patient demographics, length of stay before blood culture collection, presence of central line, vital signs, laboratory results, and SIRS and qSOFA scores [20]. They employed various models including NN and LR, with their best models achieving modest performance metrics (highest specificity at 89% but with low sensitivity). This study extensively used vital signs and other real-time clinical parameters such as temperature and heart rate as predictors. Despite this, the highest sensitivity achieved was only 31% with Logistic Regression, and even though some of their models achieved high specificity, they struggled with very low sensitivity, limiting their practical utility in clinical settings. Our model’s ability to outperform these studies without the need for immediate clinical data or vital signs not only showcases the robustness and efficiency of our approach but also suggests a potential for earlier and simpler implementation in clinical workflows. This is particularly advantageous in healthcare settings where immediate comprehensive data collection is challenging, offering a powerful tool for early BSI detection that is less dependent on the specific timing of clinical data acquisition. Similar to our study, the study by Garnica et al. (2021) used SVM, RF, and K-nearest neighbors (KNN), with a combination of RF and SVM yielding the most robust performance metrics [21]. The number of days in ICU before BC extraction, presence of catheters, age, chronic respiratory disease, fever, and CRP were reported as key predictors. Their better perfomance in comparison can also be attributed to the fact that they used a BSI dataset with prevalence rate of 51.3% which is highest reported among all the studies reporting ML based BSI prediction models.

Our study’s findings also contribute to the evolving landscape of ML applications for BSI prediction in ED settings. Schinkel et al. employed LR and XGBoost models using vital signs, laboratory results, and demographics [35]. Their XGBoost model achieved an AUROC of 0.81 (95% CI 0.78-0.83), slightly higher compared to our XGBoost model AUROC of 0.7951 (95% CI 0.7833-0.8069) and an AUPRC of 0.34 (95% CI 0.29-0.38) slightly lower than our XGBoost model AUPRC of 0.3942. The key predictors brought out by this study were temperature, creatinine, CRP, lymphocytes, DBP, bilirubin, thrombocytes, neutrophils, APL, heart rate, SBP, leukocytes, glucose, age, potassium, BUN, sodium, monocytes. Their model effectively reduced unnecessary blood cultures by approximately 30% during real-time prospective evaluation, which aligns with our goals of enhancing diagnostic efficiency and reducing healthcare costs. Our study builds on this foundation by implementing a similar ML approach but extends its application by incorporating a wider array of clinical variables derived from historical EHRs and employing a novel algorithmic configuration that may provide improved predictive performance. Similar to our study, Boerman et al. focused on the ED setting of a large teaching hospital, developing predictive models specifically for BSI outcomes based on data available at the end of ED visits, such as demographics, vital signs, administered medications, and laboratory results. They reported an AUC of 0.77 for their gradient-boosted tree (GBT) model and 0.78 for their LR model, indicating good performance in predicting bacteremia in ED [33]. The key predictors in their models were bilirubin, urea, lymphocytes, pulse rate, CRP, neutrophil, age, Temperature, DBP, potassium, glucose, thrombocytes, creatinine, ALP, SBP, and organ damage. The study highlighted the ability of their models to significantly reduce unnecessary blood cultures by predicting negative outcomes with a high degree of accuracy, reflected by a negative predictive value of over 94%. In parallel, our model not only confirms these findings but also demonstrates improved predictive accuracy and efficiency in differentiating between positive and negative BSI outcomes, which could further optimize the use of resources in ED settings. In another study, Choi et al. (2022) demonstrated the effectiveness of XGBoost model, to predict bacteremia at different stages of patient care in the ED, achieving an AUROC up to 0.853 [31]. The key predictors in the study were chief complaint, age, temperature, heart rate, and DBP at triage stage, and neutrophils, platelets, CRP, chief complaint, and creatinine at disposition stage. Their phased approach using predictions at both triage and disposition stages aligns with our methodology of employing dynamic modeling to adapt predictions based on real-time data updates. However, our model differentiates itself by utilizing a more complex array of inputs and a refined computational technique that enhances predictive generalizability. By extending the predictive capabilities beyond structured data, our study underscores the importance of comprehensive data utilization in developing models that can more effectively guide early clinical decisions in real-time settings. Our model’s ability to accommodate a broader array of clinical variables may explain any improvements in prediction robustness compared to the framework used by Choi et. al. (2023) [32]. The key predictors in their study were age, vital signs, history of chills, ambulance use. The collective insights from these comparisons suggest that while our model shares common ground with existing approaches, it also explores additional layers of complexity such as patient recent hospital interactions, comorbidities, and previous history of infections, which may influence the generalizability and effectiveness of the model across diverse healthcare environments, capturing more subtle nuances of BSI risk factors and could be pivotal in reducing BSI misdiagnosis.

Our framework’s modular design facilitates the inclusion of additional data types as they become available, enhancing its adaptability across various clinical settings. This feature is particularly valuable in current healthcare environments where flexibility and comprehensive data utilization are crucial for advancing diagnostic accuracies. Moreover, the intuitive nature of the XBSI framework ensures that it can be seamlessly integrated into existing clinical workflows, making it a practical tool for clinicians seeking to leverage AI for improved patient outcomes. The principal clinical value of our approach lies in the ability to identify patients at low risk of a positive BC, at the time of suspicion of BSI without the need for waiting to capture vital signs at the moment or within a specified time window, which could increase patient risk and stay [53, 54]. Integrating our proposed framework in hospitals as a pre-emptive BSI prediction tool can reduce BC ordering and its resulting costs and harms [49]. As it is reported in the literature that the use of data that is not routinely captured in clinical practice is the main reason why none of the prediction models have been implemented in the clinical practice yet [50]. Moreover, in the context of ICU patient monitoring, the application of AI to real-time data may seem redundant. Patients in critical care are already under intensive surveillance, and the utilization of AI for immediate alert systems could potentially clutter the workflow rather than enhance it. Instead, a more strategic use of AI lies in its ability to predict a patient’s worsening condition well before critical thresholds are reached. By analyzing historical EHRs, AI can identify subtle patterns and indicators of decline that precede acute episodes, thereby enabling preemptive medical interventions. Moreover, as the complexity of predicting health events increases the earlier the prediction occurs, achieving perfect accuracy, as often highlighted through metrics like AUC in ICU settings [38–43], may not be as critical as maintaining reasonable predictability at the initial stages of patient contact. This approach could shift the focus from crisis response to proactive patient care management, optimizing outcomes through early and targeted intervention. The only limitation of this study is its reliance on data from a single center, which may not represent the diverse patient demographics. Despite these constraints, our model’s ability to integrate a broader spectrum of data and apply a real-time learning framework supports its potential utility in clinical settings, promising reductions in unnecessary interventions and improvements in patient management. Furthermore, our use of historical patient data and variety of ML techniques allowed for a more flexible and scalable prediction framework. This aspect of our model enhances its applicability and provides a significant contribution towards advancing BSI prediction using AI.

## 5. CONCLUSIONS

This study presents a simple and intuitive XAI framework which comprehensively captures the complete medical history of a patient to accurately predict the risk of BSIs. The XBSI framework is the first of its kind to significantly enhances the predictive analysis by integrating information stored as diagnostic and procedural ICD-10 codes. This novel approach diverges significantly from traditional real-time monitoring systems, emphasizing incorporating comprehensive historical patient data. The key predictors include the count of ICD-10 codes for infectious diseases, and kidney and urinary disorders, total bilirubin, creatinine, leukocytes, thrombocytes, age, prior positive blood culture tests, and the total length of hospital stay till the blood culture test. To further enhance the generalizability of our findings and adoption of our framework, there is a need to validate our framework on administrative datasets of hospitals outside Norway and include more diverse data sources, such as biomarkers, genotypes, and phenotypes in future works.

## Data Availability

The jupyter notebooks implementing the data preprocessing, sequence creation, feature engineering and model development pipeline are available through the corresponding author. The processed and transformed final datasets, and the list of derived features are available through the corresponding author on a reasonable request.

https://docs.google.com/document/d/1yd2b4iFS8B1Vr8oc7n1Cn7iccFqZNHJo/edit?usp=drive_link&ouid=111476468840025256769&rtpof=true&sd=true

## Acknowledgements

We would like to thank the researchers at Mid-Norway Centre for Sepsis Research for valuable discussions and feedback.

## Author’s contributions

RB conceptualized and designed the study, with inputs from ØN, JEA, BE and JKD. ØN and LGT secured the data and funding for this project. RB performed data preprocessing, model development and interpretation. RB wrote the initial draft of the paper, to which the remaining authors provided comments and feedback. All authors reviewed and approved the final manuscript.

## Ethics

The use of the EPRs data in this project has been approved by the Regional Commitees for Medical and Health Research Ethics (REK) in Central Norway by REK no. 2020/26184.

---

**Table 1.** The key characteristics of the studies using ML for BSI prediction.

| Setting | First author, year | Target condition | No. Patients | Data source | Prevalence | ML models | Key predictors |
| --- | --- | --- | --- | --- | --- | --- | --- |
| Inpatients | Bhavani et al. (2020) [14] | Bacteremia and Fungemia | 76688 | EHRs, University of Chicago Hospital (2008-2018) and Loyola University Medical Center, USA (2007-2017) | Bacteremia 7.7%, Fungemia 0.7% | LR, GBM | Time from admission to BC, Temperature, Age, HR, Prior Bacteremia/Fungemia, WBC, BUN, Glucose, DBP, SBP, PPI, RR |
|  | Lee et al. (2019) [15] | Bacteremia | 13402 | EHRs, Gangnam Severance Hospital, Seoul, Republic of Korea, (2008-2012) | 7.9% | MLP, SVM, RF | ALP, PLT, Temperature, SBP, WBC, ICU stay, CRP, CVC, Age, PT, Hospital days to BC, HR, Gender, Antibiotics, RR, Creatinine |
|  | Lee et al. (2022) [16] | Bacteremia | 622771 samples | EHRs, Sinchon and Gangnam Severance Hospitals, Republic of Korea, (2007-2018) | 6.2% | MLP, RF, XGB | PLT, Monocyte, Neutrophil, Bilirubin, Albumin, and Hospital stay, BUN, ALP, RR, PR, DBP, TP, WBC, PT, Hb, CRP, Creatinine, ALT, AST, Sodium, Chloride, ESR |
|  | Cheng et al. (2020) [17] | Bacteremia | 28043 | EHRs, Zhengzhou University Hospital, China, (2017-2018) | 10% | LR, NB, SVM, ADT, CNN, BiLSTM, ABiLSTM+ DAE | Textual chief complaints, Admission records, and Laboratory biochemical indicators. |
|  | McFadden et al. (2023) [18] | Bacteremia | 10965 samples | CBC/DC, CPD, Sir Charles Gairdner Hospital, Western Australia (2018-2020) | 7.58% | RF, XGB | CBC, DIFF, and CPD |
|  | Lien et al. (2022) [19] | Bacteremia | 366586 samples | EHRs, CBC/DC, Linkou Chang Gung Memorial Hospital (CGMH) in Taiwan, (2014-2019) | 8.2% | RF, LR | CBC/DC, CRP, and PCT |
|  | Mahmoud et al. (2021) [20] | Bacteremia | 7157 | EHRs, King Abdulaziz Medical City, Riyadh, Saudi Arabia (2017-2019) | 11.4% | NN, RF, LR, DT, NB, SVM | Age, Antibiotics use, Surgery within 14 days, CVC, length of hospitalization before BC, RR, SBP, Temperature, DBP, HR, WBC, Sodium, PLT, Albumin, Creatinine, Lactic acid level. |
|  | Garnica et al. (2021) [21] | Bacteremia | 4357 | EHRs, Microbiological data, Hospital Universitario de Fuenlabrada, Madrid, Spain, (2005-2015) | 51.3% | SVM, RF, KNN | The number of days in ICU before BC extraction, presence of Catheters, Chronic Respiratory disease, Fever, Age, CRP, PLT. |
|  | Murri et al. (2024) [22] | HA-BSI | 5660 samples | Generator Center at the Fondazione Policlinico Universitario A. Gemelli IRCCS (FPG), Rome, Italy (2016-2019) | 33.6% | LR | Time BSI > 12 days, Procalcitonin > 1 ng/mL, Presence of a CVC, PLT, Hypotension, BUN, Presence of urinary catheter, Fever, Tachycardia, Altered mental status, Age, Bilirubin, Creatinine |
| Inpatients with SIRS | Ratzinger et al. (2018) [23] | Bacteremia | 466 | EHRs Vienna General Hospital, Austria, (2011-2012) | 28.8% | RF, ANN, ENR | PCT, LBP, Albumin, Bilirubin |
| Inpatients with CVC | Rahmani et al. (2022) [24] | CLABSI | 27619 | EHRs, a proprietary national longitudinal EHR repository, Houston, Texas, USA (2015-2020) | 1% | XGB, DT, LR | Temperature, HGB, comorbidities, Age, WBC, Race, Neutrophil. |
|  | Beeler et al. (2018) [25] | CLABSI | 70218 | EHRs, Indiana University Health Academic Health Center, USA, (2013-2016) | 0.6% | RF, LR | Age, Gender, history of CLABSI, CHG (Chlorhexidine Gluconate) Bathing Non-compliant Days, Line days. |
| HD patients | Zhou et al. (2023) [26] | Bacteremia | 391 | EHRs, Department of Nephrology, Affiliated Hospital of North Sichuan Medical College, Sichuan Province, China, (2018-2022) | 18.9% | LR, SVM, DT, RF, XGB | PCT, Temperature, Non-arteriovenous fistula dialysis access, NLR, Leukocyte, dialysis duration, LMR, Albumin, Neutrophil, PLT, Age, DBP, CRP, PLR, ALP, SBP, HR, history of BSI, |
| Cancer patients | Yoo et al. (2021) [27] | Candidemia | 34574 | EHRs, academic single hospital in Seoul, Republic of Korea, (2010-2018) | 0.6% | LR, ANN, RF, GBM, AML | Variables reflecting the dynamic status of patients with cancer, including blood urea nitrogen level, 7-day variance of RR, Total bilirubin level, 7-day variance of SBP, Body weight. |
| Maternity patients | Mooney et al. (2020) [28] | Bacteremia | 129 | CBC parameters, Rotunda Hospital, Ireland (2019) | 3% | CART, LDA, KNN, SVM, RF | NLR, CBC parameters. |
| Patients with PCT $\leq 2.0$ ng/ml | Su et al. (2021) [29] | Bacteremia | 931 | EHRs, Mindong Hospital Affiliated to Fujian Medical University, China, (2014-2020) | 47% | ANN, KNN, LR, RF, SVM, and NB. | Interleukin-6, PCT, D-dimer, Lactic acid, Leukocytes, Neutrophil, and PLT. |
| HIV patients | Wu et al. (2023) [30] | Bacteremia | 498 | EHRs, Wenzhou Central Hospital, China, (2014-2021) | 34.3% | SVM, ANN, GBM, GLM, MDA, PLR, NB, RF | Low Hb, CD4+T cell, PLT, LDH, BUN, splenomegaly, absence of ART treatment, Strip shadow, Nodular shadow, and Shock. |
| ED | Choi et al. (2022) [31] | Bacteremia | 24768 | EHRs, An urban tertiary referral hospital, Republic of Korea, (2016-2018) | 12% | XGB, RF, LR | Chief complaint, Age, Temperature, HR, and DBP at triage stage. Neutrophils, PLT, CRP, Chief complaints, and Creatinine at disposition stage. |
|  | Choi et al. (2023) [32] | Bacteremia | 15362 | EHRs, Seoul National University Hospital, Seoul National University Bundang Hospital, Republic of Korea, (2016-2018) | 10.9% | BNN | Age, HR, Temperature, DBP, History of chills, Ambulance use |
|  | Boerman et al. (2022) [33] | Bacteremia | 4885 | EHRs, Amsterdam UMC, location VU University Medical Center, NL, (2018-2020) | 12.2% | GBT, LR | Bilirubin, Urea, lymphocyte, Pulse rate, CRP, Neutrophil, age, Temperature, DBP, Potassium, Glucose, Thrombocytes, Creatinine, ALP, SBP, Organ damage |
|  | Chang et al. (2023) [34] | Bacteremia | 20636 | EHRs, CPD, CBC/DC, China Medical University Hospital, Taiwan, (2021-2022) | 10.4% | CatBoost, LGBM, XGB, RF, LR | Demographics, CPD, CBC/DC |
|  | Schinkel et al. (2022) [35] | Bacteremia | 6421 | EHRs, Amsterdam UMC, (VUMC, AMC, ZMC, and BIDMC), NL, (2016-2021) | 5.4% - 12.3% | XGB, LR | Temperature, Creatinine, CRP, Lymphocytes, DBP, Bilirubin, Thrombocytes, Neutrophils, ALP, HR, SBP, Leukocytes, Glucose, Age, Potassium, BUN, Sodium, monocytes |
| ED patients with SIRS | Goh et al. (2022) [36] | Bacteremia | 40395 | EHRs, National Cheng Kung University Hospital, Taiwan, (2015-2019) | 10% | LR, SVM, RF | Age, Gender, COPD, Uncomplicated DM, Hemato-oncology, WBC, Band cell, Platelet, Temperature, HR, mild liver disease, Mean arterial pressure, RR, GSC |
| ED patients<br>with fever | Tsai et al. (2023)<br>[37] | Bacteremia | 3669 | EHRs, Chi Mei Medical Center, Taiwan,<br>(2017-2020) | 13.8% | RF, LR, MLP, XGB,<br>LGBM | Hypertension, Gender, Temperature, DM, Age, CRP, PLT, WBC,<br>Malignancy, Eosinophil, HR, BMI, Hb, RR, SBP, DBP, Band, CKD,<br>Liver Cirrhosis, COPD, GCS |
| ICU<br>patients | Roimi et al.<br>(2020) [38] | Bacteremia | 3372 | EHRs, BIDMC, Boston, Massachusetts,<br>USA, (2008-2012), ICU of Rambam<br>Healthcare Campus (RHCC), Israel, (2013-2017) | ICU<br>acquired:<br>6.4%<br>(BIDMC),<br>15.9%<br>(RHCC) | RF, XGB | Time duration (days) between sampling time and last defecation,<br>Time duration (hours) between sampling time and the maximum<br>BUN (mg/dL) value measured during the 5 days prior to sampling,<br>Length of stay (days) between sampling time and ICU admission,<br>The minimal weight (kg) during the 5 days prior to sampling, The<br>time duration between sampling time and the maximum MCHC<br>(g/dL) during the 5 days prior to sampling |
|  | Van Steenkiste et<br>al. (2019) [39] | Bacteremia | 2177 | EHRs, ICU, Ghent University Hospital,<br>Belgium, (2013-2015) | 10.5% | BiLSTM, ANN,<br>SVM, KNN, LR | Temperature, Thrombocytes, Leukocytes, CRP, sepsis-related organ<br>failure assessment, HR, RR, PT, and mean systemic arterial pressure. |
|  | Boner et al.<br>(2022) [40] | Bacteremia | 6557 | EHRs, ICU, University of Virginia, USA,<br>(2011-2015) | 13.3% | FNN, GRU, CNN,<br>LR | Temperature, BUN, BP, HR, Albumin, PLT, Chloride, Creatinine,<br>Chloride, and Phosphorus. |
|  | Pai et al. (2021)<br>[41] | Bacteremia | 4275 | EHRs, Taichung Veterans General Hospital<br>ICU, Taiwan, (2015-2019) | 13.8% | LR, SVM, MLP, RF,<br>XGB | ALP, CVC period, prothrombin time, PLT, Albumin, Apache II<br>score, Age, foley |
| ICU<br>patients<br>with CVC | Parreco et al.<br>(2018) [42] | CLABSI | 57786<br>admissio<br>ns | MIMIC-III database, USA, (2001-2012) | 1.5% | LR, GBT, DL | Severity of illness scores (like SAPS II, APS III, and OASIS) and<br>comorbidities. |
| ICU<br>patients<br>with new-<br>onset SIRS | Yuan et al. (2021)<br>[43] | Candidemia | 7932 | EHRs, Peking Union Medical College<br>Hospital, The Affiliated Hospital of Qingdao<br>University, The First Affiliated Hospital of<br>Fujian Medical University, China, (2013-2017) | 1% | XGB, SVM, RF, ET,<br>LR | Colonization, Diabetes, AKI, total number of parenteral nutrition<br>days, history of fungal infection, CRRT days, Abdominal surgery,<br>BDG, days of mechanical ventilation, Length of hospital and ICU<br>stay, days of CVC |

